# Investigating the prevalence of tuberculosis and associated risk factors among prisoners at Peter Singogo Prison in Ndola: a retrospective cross-sectional study

**DOI:** 10.1101/2024.05.01.24305425

**Authors:** Chipo Manda, Namaunga Kasumu Chisompola

**Affiliations:** Department of Basic Medical Sciences, Michael Chilufya Sata School of Medicine, Copperbelt University, Ndola, Zambia

**Keywords:** Prevalence, Tuberculosis (TB), susceptibility, risk factors, prisoners

## Abstract

**Background:** Tuberculosis primarily affects the lungs and is influenced by risk factors such as diabetes, HIV/AIDS, substance abuse, and crowded living conditions. Prisons can be high-risk environments for tuberculosis transmission due to overcrowding, poor ventilation, and limited access to medical care. Substance misuse and HIV infection are common among prisoners, increasing the risk of tuberculosis. The Peter Singogo Prison in Ndola, Zambia was studied to determine the prevalence of tuberculosis and its associated risk factors.

**Methods:** To ascertain the prevalence of Tuberculosis and related risk factors among inmates at Peter Singogo Prison, a retrospective cross-sectional study was done. Out of the 450 prisoners who passed screening, 220 prisoners were chosen using a purposive sampling strategy after Tuberculosis records were reviewed, considering known risk factors, Tuberculosis symptoms, and Sputum microbiology laboratory results. The collected results were then examined using Statistical Package for the Social Sciences Version 23; statistical significance was recognized for a P value of less than or equal to 0.05.

**Results:** Two hundred and twenty prisoners had Tuberculosis symptoms. From these, a total of 51 prisoners (23%) had a Tuberculosis diagnoses with only 18 prisoners (35%) having a bacteriological confirmation of Tuberculosis, while 33 prisoners (65%) had a clinical diagnosis. In cases with bacteriological confirmation, the smear was positive. Human Immunodeficiency Virus positive inmates had a much greater frequency of Tuberculosis than Human Immunodeficiency Virus negative inmates (80% vs. 20%).

**Conclusion:** A significant prevalence of Tuberculosis at Peter Singogo prison was identified, with the majority of cases occurring in inmates who were Human Immunodeficiency Virus positive. The high prevalence found in this study may help to explain how Tuberculosis spreads within the prison, amongst inmates, and among different communities. Therefore, it is important to pay attention to prison environments to stop the spread of Tuberculosis to both convicts and the wider public.

## Introduction

Tuberculosis (TB) is a highly infectious illness caused by Mycobacterium tuberculosis (Mtb), which usually affects the lungs to form pulmonary TB and other body areas to cause extrapulmonary TB [1]. Airborne droplets created by coughing, sneezing, or talking are inhaled by people in touch [1, 2].

Tuberculosis is a major global health problem which affects about 30% of the world’s population and is the topmost cause of mortality from a sole infectious agent accounting for more than 1.3 million deaths yearly [1,2].

There are significant concerns in two parts of the world: Africa where the Human Immunodeficiency Virus (HIV) infection is also prevalent and Eastern Europe. The situation in Eastern Europe is critical due to multi- drug resistant (MDR) and extensively drug-resistant (XDR) types of tuberculosis. However, recent trends pose concerns on the rising prevalence of drug resistant forms of TB in Africa too [3]. The World Health Organization’s south-east Asia and Africa areas had the largest number of TB cases and disease incidence, with HIV infection accounting for 9% of cases [1]. It is also estimated that appropriately 94% of all TB infections and deaths occur in low- and-middle-income countries, including Sub-Saharan Africa (SSA) [1, 3].

Inmates or convicts are categorized as TB important populations with a high risk of exposure to MTB and getting TB infection, yet they are frequently overlooked for a variety of established reasons [4, 5]. In comparison to the general population, research has shown that people in prison have a higher chance of contracting and developing tuberculosis [6, 7]. Overcrowding, poor ventilation, poor nutrition, concomitant illnesses such as HIV, and a lack of access to proper health care services are typical causes, particularly in jails in (SSA) nations; which house a considerable number of people living with HIV or at risk of contracting the virus [8].

They include people from impoverished communities, those with poor baseline health, and those who engage in high-risk behaviors, such as commercial sex workers or individuals with substance use disorders.

As a result, incarcerated populations in SSA correctional facilities, who encompass both the sentenced offenders and those detained and awaiting trial (on remand), experience higher HIV and TB prevalence than the general population [9, 10]

Studies from Zambia, a country among those having the largest correctional populations and generalized HIV epidemics in Southern Africa, has reported HIV prevalence among incarcerated persons between 12.5% and 27.4%, and TB prevalence of 0.34% to 7.6 [10]. Hence, there is the need to sustain active search for TB cases in Zambian prisons. Tuberculosis active case finding in prisons may involve approaches such as mass screening, entry screening, routine screening, and exit screening. Active case finding of TB cases contributes to increasing TB case notification and treatment success rates and reducing mortality [11].

The prevalence of TB in the patients with a history of imprisonment is not known in the region. Tuberculosis remains a major public health concern, exacerbated by the delay and poor quality of diagnosis combined with a deficient healthcare system, particularly in the prison environment [11]. Pulmonary tuberculosis contact investigations are rarely and inconsistently carried out in resource-limited settings of low- and middle-income countries like Zambia. Zambia ranks 13th among countries with the highest TB burden in the world and it is estimated that 6.3% of TB cases in Zambia are attributable to TB transmission within correctional [12]. There is a high rate of pulmonary TB in Zambian prisons, with significant rates of drug resistance and MDR-TB, highlighting the need for active surveillance and treatment programs [13]. However, the National Tuberculosis and Leprosy Program (NTLP) currently does not have a robust recording and reporting mechanism on TB case detection and treatment outcomes from correctional facilities [14, 15].

Prisoners have a high occurrence of conditions that increase the risk of TB, including substance abuse and HIV infection. Prisons act as infectious disease reservoirs. The increased prevalence of communicable diseases among people in prison can constitute a risk for the health of people who live/work in prison settings and for the general population, as the vast majority of people in prison eventually return to their communities [16]. There are several risk factors associated with increased transmission rates in prison settings, e.g. proximity (aggravated by overcrowding), which is common in nearly all Zambian correctional facilities; high-risk sexual behavior; injecting drug use; sharing of injecting equipment; tattooing and piercing [16, 7]. The study aimed to determine the prevalence of TB and its associated risk factors at Peter Singogo Prison in Ndola, Zambia; to further elucidate the extent of the TB burden in prisons in Zambia [17].

## Methods

### Study site

Peter Singogo prison is a medium security facility built in the mid 1930’s by Northern Rhodesia. It has a capacity of 450 with 200 inmates being juvenile and 350 being adults; among which is a section for the convicted and another section for those on remand with less upgrade in the infrastructure. The facility is located at the center of mid-densely populated services which includes Ndola Teaching Hospital and surrounded by several low-mid income residential compounds.

### Study design

A retrospective cross-sectional study was used to determine the prevalence of TB and associated risk factors among prisoners at Peter Singogo prison. Data was collected retrospectively from records between March 2021 and March 2022.

### Sample size and technique

Purposive sampling technique was used to select 51 prisoners from records of inmates that met the inclusion criteria. The prisoners included in this study consisted of inmates and staff members presenting with signs and symptoms of TB, those that did not have any symptoms of TB were not included. Data collection: A retrospective cross-sectional study was conducted to determine the prevalence of TB and associated risk factors among prisoners at Peter Singogo prison. TB records were reviewed, and 220 Prisoners were selected using purposive sampling technique based on TB Symptom, reported risk factors and Sputum microbiology laboratory results. A questionnaire was used as a guide to purposely select the participants’ records, a TB case was defined from bacteriological records (culture results/smear) or clinical records (symptoms) at Ndola Teaching Hospital pathology department. Thereafter, the information obtained was processed using the statistical package for social sciences version 23 using the chi square with level of significance accepted at values equal or less than 0.005 at 95% confidence interval.

### Ethical Considerations

Ethical consideration for the study was obtained from Tropical Disease Research Centre (TDRC) ethics committee after the submission and review of the study protocol, study IRB number: 00002911. Additional permission was obtained from the Ndola district health office. The need for consent was waived by the Tropical Diseases Research Centre Ethics Committe because the study was a retrospective study and some of the records obtained were for prisoners who had been released or transferred at the time of the study. In this study, it was necessary to trace laboratory results back to the patient’s clinical data. As a result, complete anonymity could not be maintained. However, all patient records were stored securely, and access was restricted to only the research staff involved in the study period. This measure was taken to ensure the confidentiality and privacy of the patient’s information while still allowing for the necessary data linkage required for the research.

## Results

Symptoms were found in 220 (48%) prisoners out of the 450 screened inmate’s s). However, TB was diagnosed in 51 individuals representing a prevalence of 23 % from records that matched with the inclusion criteria and had one or more of the World Health Organization (WHO) recommended screening symptoms of cough, fever, weight loss or night sweats, of which cough [84(38%)] and night sweats [78(35%)] were commonly recorded (Table 1).

**Table 1:**
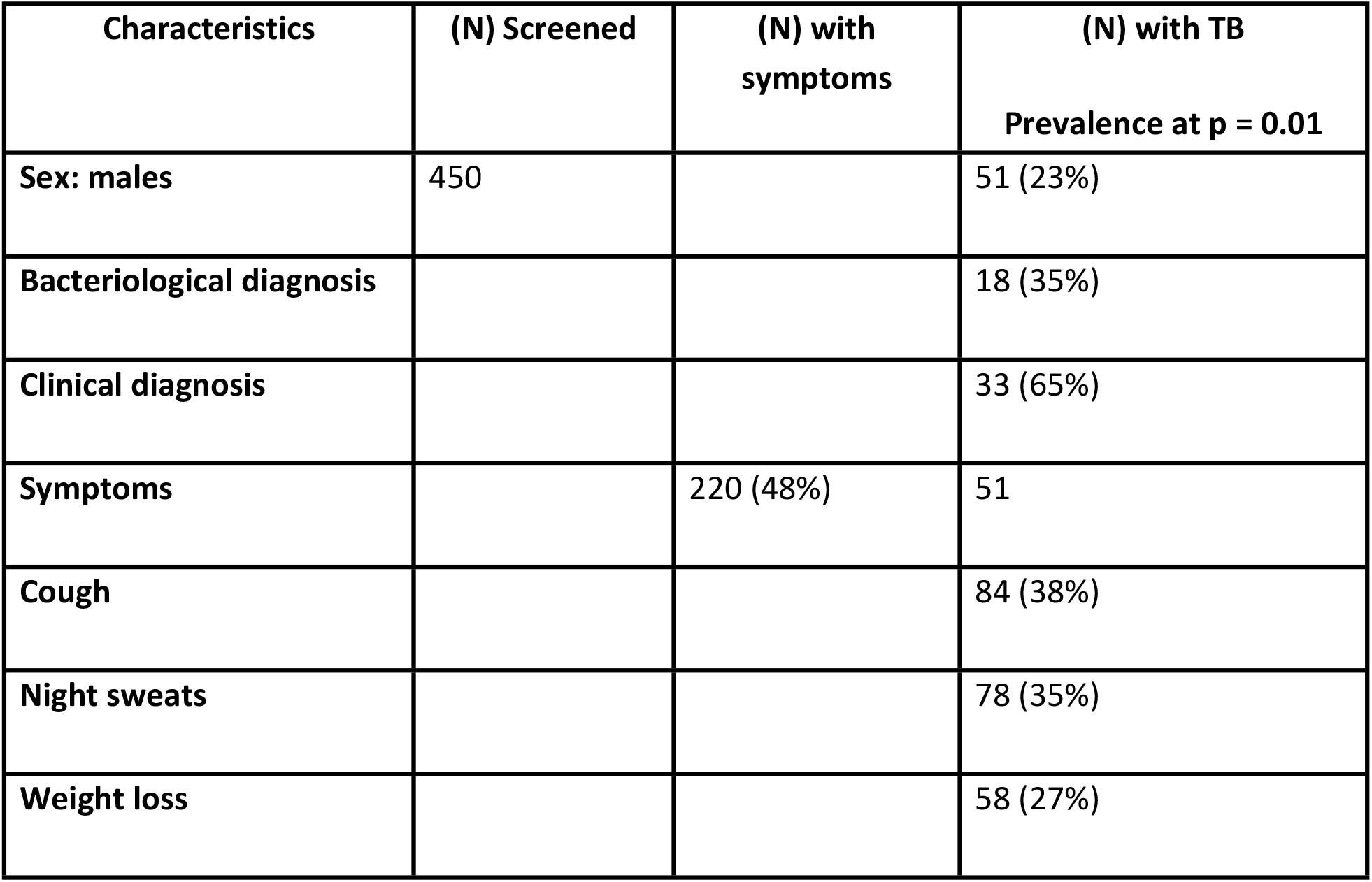
Prevalence and population characteristics.

| Characteristics | (N) Screened | (N) with symptoms | (N) with TB<br>Prevalence at p = 0.01 |
| --- | --- | --- | --- |
| Sex: males | 450 |  | 51 (23%) |
| Bacteriological diagnosis |  |  | 18 (35%) |
| Clinical diagnosis |  |  | 33 (65%) |
| Symptoms |  | 220 (48%) | 51 |
| Cough |  |  | 84 (38%) |
| Night sweats |  |  | 78 (35%) |
| Weight loss |  |  | 58 (27%) |

Tuberculosis was bacteriologically confirmed by smear microscopy in 18 cases (35%) and clinically diagnosed in 33 cases (65%). TB prevalence was significantly higher in HIV positive inmates [9% (41), 0.01] than in HIV negative inmates [2% (10), 0.01], as demonstrated in Table 2.

**Table 2:** HIV prevalence and TB/HIV co-infection.

| Proportion of HIV Positive inmates with TB | Proportion of HIV negative inmates with TB | P value |
| --- | --- | --- |
| 41(80%) | 10(20%) | 0.01 |

## Discussion

This study investigated the prevalence of TB among prisoners at Peter Singogo prison. The study found a TB prevalence among prisoners of 23% (p=0.01) representing a total of 51 inmates bacteriologically (35%) and clinically(65%) diagnosed with TB. According to the World Health Organization’s Global Tuberculosis Report 2021, the estimated national prevalence of TB in Zambia’s general population was 392 cases per 100,000 population (0.392%) in 2020. This means that approximately 70,000 people were living with TB in Zambia in 2020. The TB prevalence among prisoners of 23% found in this study is significantly higher that of the national prevalence of TB in Zambia in 2020, which was approximately 0.392% [1].

A number of prison studies have reported on various prevalence of TB as well as HIV/TB prevalence among inmates. The knowledge of these studies in sub-Saharan African prisons reports presents baseline prevalence for both TB and TB/HIV co-infection [6]. In view of this well-established biological as well as clinical linkage between TB and HIV, the high baseline rates of infection and the acknowledged high-risk environment of prisons, hence, these findings constitute an important contribution to the evidence base.

The findings of this study are similar to most studies conducted elsewhere; a study in Ethiopia estimated the prevalence of TB among prisoners to be 8.33% at 95% CI and p < 0.001 [15, 14]. All TB diagnosis relied on microscopy which has low detection rate and 50% of the included studies used microscopy as a confirmation of TB. Alternatively, rapid molecular techniques such as the GeneXpert, can be introduced in prisons for detection of TB and rifampicin resistance. In addition, screening was solely dependent on symptoms of cough [12]. This pooled prevalence is comparable with reports from South African prisons with a prevalence rate of 8.8% [9]. On the other hand, Democratic Republic of Congo reported a high TB prevalence of 17.7% compared to the prevalence in this study (23%) which was slightly high [9]. This might be due to variations in the laboratory detection methods used to diagnose TB and can also be attributed to missed cases. In another study conducted in Lusaka central prison, TB prevalence was found to be 23% which was high and same as the findings of this study against a prevalence of 13% for the whole Lusaka province [15, 14] This high prevalence was also comparable to a study in Malawi whose prevalence was found to be 25% [9]. This high disease burden may be attributed to the poor health conditions of prisons which affects the health of inmates and may further threaten the health of the community.

TB in this study was bacteriologically confirmed in 33 cases (65%) and clinically diagnosed in 18 cases (35%). Smear was positive in bacteriologically confirmed cases. TB prevalence among prisoners was significantly higher in HIV positive inmates [80% (41), 0.01] than in HIV negative inmates [20% (10]. Similarly in another study, the prevalence of bacteriologically confirmed TB found in the general population of Lusaka Province together with HIV prevalence among inmates residing within Lusaka Central Prison was found to be approximately twice that of the national prevalence of 27% against 13.5% and 30% greater than in Lusaka province 27% against a prevalence of 21% (UNAIDS, 2013). These findings place emphasis on the need for routine screening and treatment for both HIV and TB in prisons in Zambia.

In addition, a study in Cameroon, found majority of prisoners diagnosed with pulmonary TB had a cough for more than 3 weeks, 14% were symptomatic for shorter periods of time (Noeske et al, 2016). Moreover, the prevalence of active TB was similar among those with cough of for more than 3 weeks against those with cough of 1 to 3 weeks, suggesting that limiting screening based on the duration of symptoms. A study of prisoners in Malawi found that 40% of cases of active TB had cough for more than 3 weeks’ duration, resulting in a change in the standard procedure for screening of inmates [9]. These findings are similar to the findings of this study where cough and night sweat were the most common features in those diagnosed with TB for more than 3 weeks duration of symptoms; demonstrating the need for early quarantine and diagnosis of symptomatic individuals. However, in some countries the pooled prevalence of TB in some prisons was very low when compared to the findings of this study and from studies cited above. A study conducted in France found TB prevalence to be 888 per 100,000 95% confidence interval [1]. Although these findings were in agreement with reports from Brazil prisons with a point prevalence of 917 per 100,000 and recent reports showed a national prevalence of 200/100,000 [1] which are several times lower than the findings of this study. Moreover, lower prevalence’s were reported from prison settings in Turkey with 195 per 100,000 [1]. The lower prevalence’s in these countries could be attributed for strong TB control strategies, low incidence rates and established good health systems both in prisons and general population. For the purpose of developing efficient diagnostic, therapeutic, and preventive approaches, additional research on a sizable prison population is required.

## Conclusion and Recommendations

The study determined a high prevalence of TB at Peter Singogo prison with most cases found in HIV positive inmates. The high prevalence of TB could explain the spread of TB within the prison and between prisoners and various communities. Thus, early case detection and treatment should be priorities in prison settings to prevent the transmission of TB both in inmates and the general population. Further studies covering large scale prison population are needed to design effective diagnostic, treatment and preventive methods.

The significant prevalence of TB and HIV in this study suggests mechanisms that may contribute to disease concentration, and transmission. These findings constitute an alert not only to the poor health conditions of prisons and inmates, but also to the way poor prisoner health may threaten community disease control efforts. To tackle this dual burden of disease, we recommend a coordinated strategy among government institutions and stakeholders needed to implement preventive measures within the prison environment. We also recommend Targeted intervention through routine and frequent HIV and TB testing in inmates.

## Data Availability

All data produced in the present work are contained in the manuscript

## Acknowledgements

The authors are grateful for the valuable suggestions made by Mespa Manyepa, Ngula Kabelenga, Paul Siyapila and Sam Miti during manuscript preparation. The authors would like to thank the staff and inmates at Peter Singogo Prison for their participation in the study.

